# A Simulation Study to Advance Human-Centred Artificial Intelligence via Digital Citizen Science: Can Large Language Models Transform Current Approaches to Missing Data Imputation?

**DOI:** 10.1101/2025.08.21.25334182

**Authors:** Jamin Patel, Keaton Banik, Sheriff Tolulope Ibrahim, Tarun Katapally

## Abstract

**Background:** Missing data is a persistent challenge in digital health research, and traditional approaches like Multiple Imputation by Chained Equations (MICE) may not capture complex patterns. While large language models (LLMs) could offer a viable alternative, their use in this context remains understudied. Moreover, a critical gap remains in embedding human-centred artificial intelligence (AI) approaches that integrate equity, transparency, and stakeholder participation. Digital citizen science, which leverages citizen-owned devices for ethical, participatory big data collection, offers a foundation to advance such approaches in digital health.

**Objective:** To evaluate and compare the imputation accuracy of MICE with the OpenAI o3 model for categorical variables in a simulated digital health dataset under different missingness mechanisms and levels, while situating this evaluation within the broader vision of human-centred AI enabled by digital citizen science.

**Methods:** A complete digital health dataset collected through a digital citizen science platform was used to simulate missingness under Missing at Random (MAR) and Missing Completely at Random (MCAR) at 10%, 25%, and 50%. MICE used logistic regression with five imputations and ten iterations per chain. For the o3 model, structured prompts were generated for each missing entry using all available non-missing variables from the same record. Both methods were evaluated on each simulated dataset using classification accuracy and a closeness metric representing similarity to the original data. Statistical differences were tested with a two-sample Z-test, and misclassification patterns were examined by variable type and category frequency.

**Results:** Under MAR conditions, MICE and o3 performed similarly with an average accuracy of 0.60 and 0.59, and closeness metrics of 0.83 and 0.85, respectively. Under MCAR, both methods achieved 0.59 accuracy, with closeness metrics of 0.84 and 0.85. No statistically significant differences were found across conditions (all p > 0.05).

**Conclusion:** While MICE remains preferred for continuous data, the o3 model shows promise as a complementary tool for categorical imputation in smaller datasets. Beyond methodological comparability, this study demonstrates how digital citizen science can serve as an ethical foundation for embedding human-centred AI into digital health research, positioning large language models not only as technical tools but also as vehicles for advancing equity, transparency, and participatory innovation in healthcare.

## 1. Introduction

As mobile devices, wearables, and app-based platforms increasingly capture high-frequency, context-rich information in clinical and community settings, the volume and complexity of available health data have expanded substantially [1,2]. These technologies enable passive sensing (e.g., continuous heart rate monitoring), real-time patient-reported outcomes (e.g., mobile ecological momentary assessments), and continuous behavioural or physiological monitoring (e.g., activity, sleep, medication adherence) across multiple domains [3–6]. Obtaining such big data consistently can enable the development of precision prediction, prevention, and management models by catalyzing the generation of artificial intelligence [AI] [7].

Digital citizen science has shown promise to ethically obtain big data from citizen-owned ubiquitous devices to advance digital health via the digital transformation of health systems [8]. Digital citizen science, a participatory approach that can range from contributory and collaborative methods (data collection and analysis) to cocreation of knowledge (conceptualization and knowledge translation), from a health systems and public health perspective, can be initiated by health system scientists to obtain longitudinal big data [9,10]. From an AI development perspective, digital citizen science can address one key gap – the embedding of participatory approaches to facilitate human-centred AI from the conceptualization stage of big data collection [11,12].

Despite these benefits, digital big data collection via ubiquitous tools is often affected by incomplete data arising from device malfunction, app crashes, sensor dropout, user nonresponse, or environmental interference [13]. Missingness is frequently Missing at Random (MAR), where the probability of missing data depends on observed participant or situational characteristics [14]. If MAR data are not addressed with appropriate methods, estimates of associations, means, or other parameters may be biased, leading to incorrect conclusions and reduced generalizability [15,16]. A common but limited approach is complete-case analysis, which excludes records with missing values [17]. This yields unbiased estimates only when missingness is Missing Completely at Random (MCAR), a condition rarely met in digital health datasets [14]. Under MAR, complete-case analysis can produce biased estimates and substantially reduce statistical power by discarding large portions of the data [18].

To mitigate these issues, researchers frequently use Multiple Imputation by Chained Equations (MICE) [19]. MICE iteratively models each variable with missingness as a function of the others, assuming MAR. When assumptions hold and models are correctly specified, MICE can produce estimates close to those from complete data. However, in high-dimensional, noisy datasets from mobile and wearable platforms, relationships may be nonlinear, involve higher-order interactions, or vary over time [20], making correct model specification challenging. While noise-aware missing data algorithms exist, they are technically complex and require substantial statistical expertise to implement [21]. MICE also requires sufficient complete cases for stability, which can be challenging in small clinical subgroups [22–24].

These constraints create an opportunity for generative AI approaches, particularly large language models (LLMs) [25]. Trained on vast, multimodal datasets that integrate language, code, and structured data, LLMs can generalize across reasoning tasks [26]. While developed for natural language processing, they can also operate on structured datasets – such as electronic health records, physiological time series from wearables, and patient-reported outcomes – when formatted as text [25,27]. By leveraging learned representations of language, logic, and statistical regularities [28], LLMs have the potential to infer plausible values for missing entries based on the broader clinical context of each patient.

Prior efforts to apply LLMs for missing data imputation in health systems research have shown promise, but many approaches require dataset-specific fine-tuning [29], which reduces scalability in heterogeneous datasets. Others depend on highly tailored prompting strategies [30], raising concerns about robustness when response options are complex or not linguistically well aligned. Architectures based on adversarial learning or feedback-augmented prompting achieved strong accuracy but relied on specialized model design, manual intervention, or access to ground truth during training [31,32]. Collectively, these studies demonstrate feasibility but have not established whether a general-purpose LLM, applied with a simple prompting strategy rather than extensive adaptation, can perform imputation in the noisy, multidimensional datasets typical of real-world digital health research.

To address current gaps in AI-assisted imputation, drawing primary data obtained via digital citizen science approaches from citizen-owned ubiquitous devices, this study compared OpenAI’s o3 model with MICE - the current statistical benchmark for handling missing data. Using a simulation framework, we aimed to evaluate the imputation performance for categorical variables under MAR and MCAR conditions commonly encountered in complex digital health data. We hypothesized that MICE would perform well under MAR, where its assumptions are satisfied, while o3 would achieve comparable accuracy across both mechanisms, providing initial evidence on the feasibility of LLMs as complementary tools for digital health imputation.

## 2. Methods

### 2.1. Study design

This simulation study drew on data collected through the Smart Platform, a digital citizen science initiative that combines community-based participatory research and systems science to engage citizens as active contributors to health systems research [33]. The Smart Platform deploys a custom-built mobile application (app) on participants’ personal smartphones to capture survey-based, behavioural, and contextual health data in real-world settings. The app facilitates secure, end-to-end encrypted data collection and enables participants to contribute information on a daily basis, thereby generating high-frequency, ecologically valid datasets suitable for digital health research.

For this analysis, we used a subset of 363 complete cases, defined as participant records with no missing values across all questionnaire items selected for this study, including demographic variables (age, gender, household income, parental education, job, and ethnicity) and mental health items (stress, anxiety, depressive symptoms, and suicidality). These complete cases provided the ground truth against which imputation performance was evaluated.

### 2.2. Participants

Participants were drawn from a larger cohort of youth and young adults aged 13-21 years recruited through secondary schools in Regina, Saskatchewan, Canada. Recruitment was conducted via in-school engagement sessions, during which the research team introduced the study, demonstrated the Smart Platform app, and supported students in downloading it onto their smartphones. Eligibility criteria required participants to be within the target age range and to have no medical conditions preventing mobile participation. Informed consent was obtained directly from participants, with implied parental consent required for those under 16 years of age. All procedures were reviewed and approved by the Research Ethics Boards of the University of Regina and the University of Saskatchewan (REB #2017-029). The present analysis was conducted on a fully de-identified dataset within a secure, sandboxed environment, with no personally identifiable information transmitted externally.

### 2.3. Measures

Fourteen categorical variables were selected to represent common features in digital health datasets (**Table 1**). These included six demographic variables (age, gender, household income, parental education, job, and ethnicity) and seven mental health survey items addressing stress, anxiety, depressive symptoms, and suicidal ideation. Response categories reflected those used in the original study instruments.

**Table 1:** Sociodemographic and mental health measures deployed via the Smart Platform with response options.

| Variable | Responses |
| --- | --- |
| In general, would you say your mental health is? | • Poor<br>• Excellent<br>• Fair<br>• Good |
|  | • Very good<br>• Don't know |
| Thinking about the amount of stress in your life, would you say that most days are: | • Very stressful<br>• Not at all stressful<br>• Extremely stressful<br>• A bit stressful<br>• Not very stressful |
| How often over the last 2 weeks were you bothered by: Feeling nervous, anxious or on edge? | • Several days<br>• Not at all<br>• Nearly every day<br>• More than half the days |
| How often over the last 2 weeks were you bothered by: Not being able to stop or control worrying? | • Several days<br>• Not at all<br>• More than half the days<br>• Nearly every day |
| During the past 12 months, did you ever feel so sad or hopeless almost every day for two weeks or more in a row that you stopped doing some usual activities? | • No<br>• Yes |
| During the past 12 months, did you ever seriously consider attempting suicide? | • No<br>• Yes |
| When was the last time that you had significant problems with Feeling very trapped, lonely, sad, blue, depressed, or hopeless about the future? | • Past month<br>• Never<br>• Over a year ago<br>• Don't know<br>• Past year<br>• Past 3 months<br>• Past 6 months |
| When was the last time that you had significant problems with Feeling very anxious, nervous, tense, scared, panicked, or like something bad was going to happen? | • Past month<br>• Never<br>• Past 3 months<br>• Don't know<br>• Past year<br>• Over a year ago<br>• Past 6 months |
| Age | • 13 years – 21 years |
| Gender | • Female<br>• Male<br>• Other<br>• Prefer not to disclose<br>• Transgender |
| Household Income | • Under \$10,000 – Over \$100,000<br>• Don't know |
|  | • Prefer not to answer |
| Parental Education | • Elementary School<br>• Some secondary/high school<br>• Completed secondary/high school<br>• Some post-secondary (university or college)<br>• Received university of college degree/diploma<br>• Do not know/does not apply |
| Job | • Yes<br>• No |
| Ethnicity | • African<br>• Asian<br>• Canadian<br>• Cree<br>• Eastern European<br>• First Nation<br>• Metis<br>• Other<br>• South Asian |

### 2.4. Missing data simulation

Missing values were introduced synthetically under both MCAR and MAR mechanisms. For MCAR, values were removed randomly and independently of other predictors. We repeated the simulation at three overall missingness levels: 10%, 25%, and 50%, with a reproducible random seed [34]. For MAR, the probability of missingness for each variable was modeled as a function of other observed features, with stochastic noise added to avoid deterministic patterns. A weak MAR structure was also generated by supplementing MAR with a small proportion of MCAR values to reflect technical failures such as app crashes or connectivity loss. Simulations were repeated at three overall missingness levels (10%, 25%, and 50%) using reproducible random seeds. The probability of missingness for variable *j* was defined as:

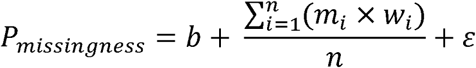

Where *n* is the number of predictors, *b* is a linear coefficient, *m* is the weight assigned to the feature *i*, *w_i_*is the normalized value of feature *i* scaled between 0 and 1, and *ε_i_ ∼ N(0,0.05^2^)* is normally distributed random noise [35,36]. For instance, if Age was modeled as missing conditional on Job (*m*=0.6, *w*=0.5) and Gender (*m* = 0.1, *w* = 0.2), the probability of Age being missing was:

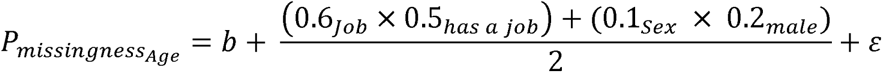

To ensure comparability across features, all data values were normalized with feature scaling between 0 and 1, and categorical variables were factorized into numeric codes prior to normalization. The normalized values functioned as predictors of missingness (*w*). Feature weights (*m*) were randomly drawn between 0.01 and 0.70, representing a 1-70% contribution to missingness probability, to mimic realistic variation in missingness drivers. The final probability was then used in a Bernoulli draw to assign missingness. This process yielded an average MAR missingness of 18.3%.

### 2.5. Imputation methodology

To compare statistical and AI-based approaches, we evaluated two methods of categorical data imputation: MICE and OpenAI’s o3 model. MICE was implemented in Python using logistic regression models for categorical variables, where each incomplete variable was imputed conditionally on all other observed variables in turn. Five imputations were generated with 100 iterations per chain, following standard practice to ensure convergence. Final imputed values were obtained by pooling across imputations and selecting the modal category.

The AI-based imputation was performed using OpenAI’s o3 model in a zero-shot setting, meaning the model was applied without any prior task-specific training or example-based guidance. For each observation with a missing value, a structured prompt was generated from the available non-missing variables. Prompts followed the format: “Given the following information about a participant in a digital health study; Age: 16, Grade: 4, Location: 2, Job: Yes, what is the most likely response to “During the past 12 months, did you ever feel so sad or hopeless almost every day for two weeks or more in a row that you stopped doing some usual activities? (1 = No, 2 = Yes)? Respond with only the number.” Each prompt was submitted independently to the o3 model through its API, and the returned response was parsed as the imputed value. No fine-tuning, few-shot examples, or ground-truth feedback were used, and the procedure was applied consistently across all missingness levels and simulation iterations.

### 2.6. Evaluating imputation performance

Imputation performance was assessed by comparing imputed values to the original complete dataset. The primary metric was classification accuracy, defined as the proportion of correctly imputed values. To account for partial agreement on ordered categorical variables, we also computed an ordinal proximity score, defined as the normalized distance between the imputed and true response, with values closer to 1 indicating greater ordinal agreement. For instance, on a five-point Likert scale (strongly disagree, disagree, neutral, agree, strongly agree), imputing “strongly agree” for a true value of “agree” would be considered closer than imputing “strongly disagree.” Responses marked “I don’t know” or “Prefer not to answer” were excluded, and binary items were not included in the ordinal proximity analysis. Statistical significance of differences in imputation accuracy between methods and across missingness levels was assessed using two-proportion z-tests, with p <0.05 considered significant.

## 3. Results

### 3.1. MICE Imputation

For the MAR dataset (n = 363), MICE imputation achieved a mean accuracy of 0.60 (SD = 0.14; range 0.35-0.81). Accuracy was lowest for “Thinking about the amount of stress in your life, would you say that most days are” (0.35) and highest for “During the past 12 months, did you ever seriously consider attempting suicide?” (0.81) (Figure 1). The mean closeness score was 0.83 (SD = 0.04; range 0.79-0.91), from 0.79 for “How often over the last 2 weeks were you bothered by: Feeling nervous, anxious or on edge?” to 0.91 for “When was the last time that you had significant problems with feeling very anxious, nervous, tense, scared, panicked, or like something bad was going to happen?”

**Figure 1:**
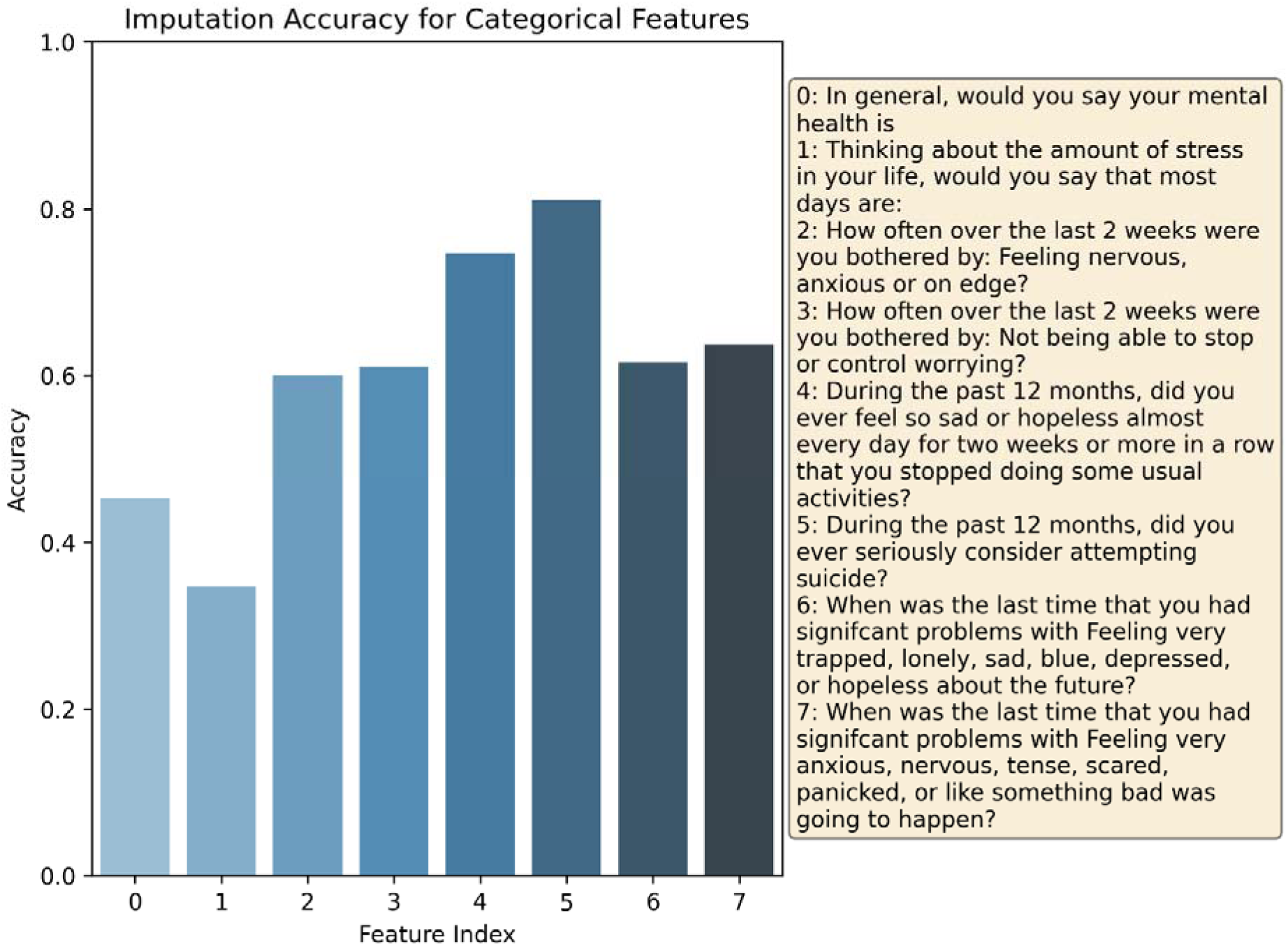
MICE Imputation accuracy from MAR generated dataset (18.32% missingness) using a normalized average to compute the probability of missingness. Each bar represents the accuracy of imputation compared to the ground truth dataset.

On the 25% MCAR dataset, MICE produced a mean accuracy of 0.59 (SD = 0.12; range 0.43-0.82), with lowest accuracy for “In general, would you say your mental health is” (0.43) and highest again for “During the past 12 months, did you ever seriously consider attempting suicide?” (0.82) (Figure 2). The mean closeness was 0.84 (SD = 0.03; range 0.80-0.88). Across both MAR and MCAR, binary questions showed slightly different misclassification patterns (Figures 3-4), and items with larger sample sizes had lower misclassification rates, as expected.

**Figure 2:**
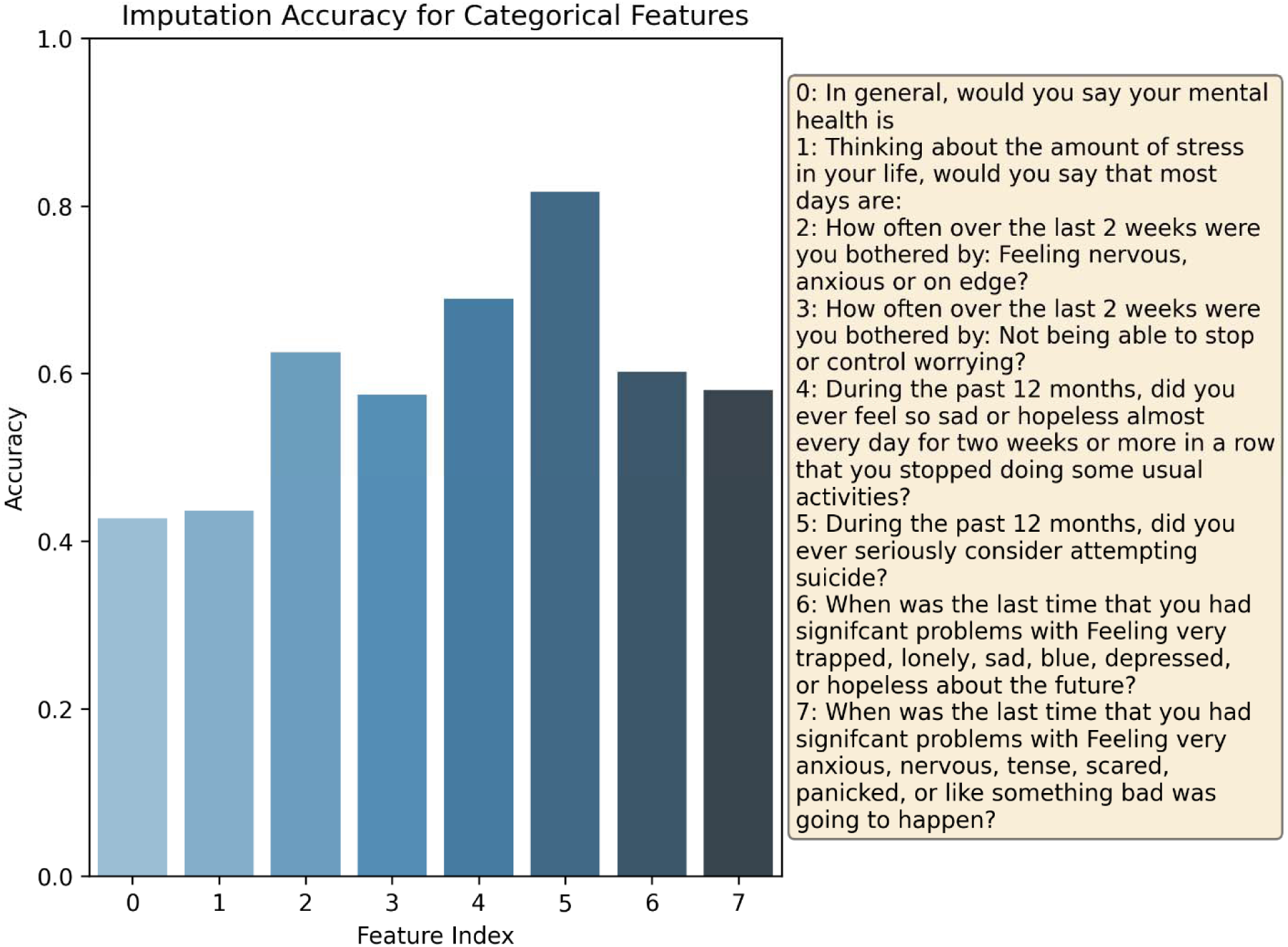
MICE Imputation accuracy from MCAR generated dataset (25% missingness) using a Bernoulli draw to determine missingness. Each bar represents the accuracy of imputation compared to the ground truth dataset.

**Figure 3:**
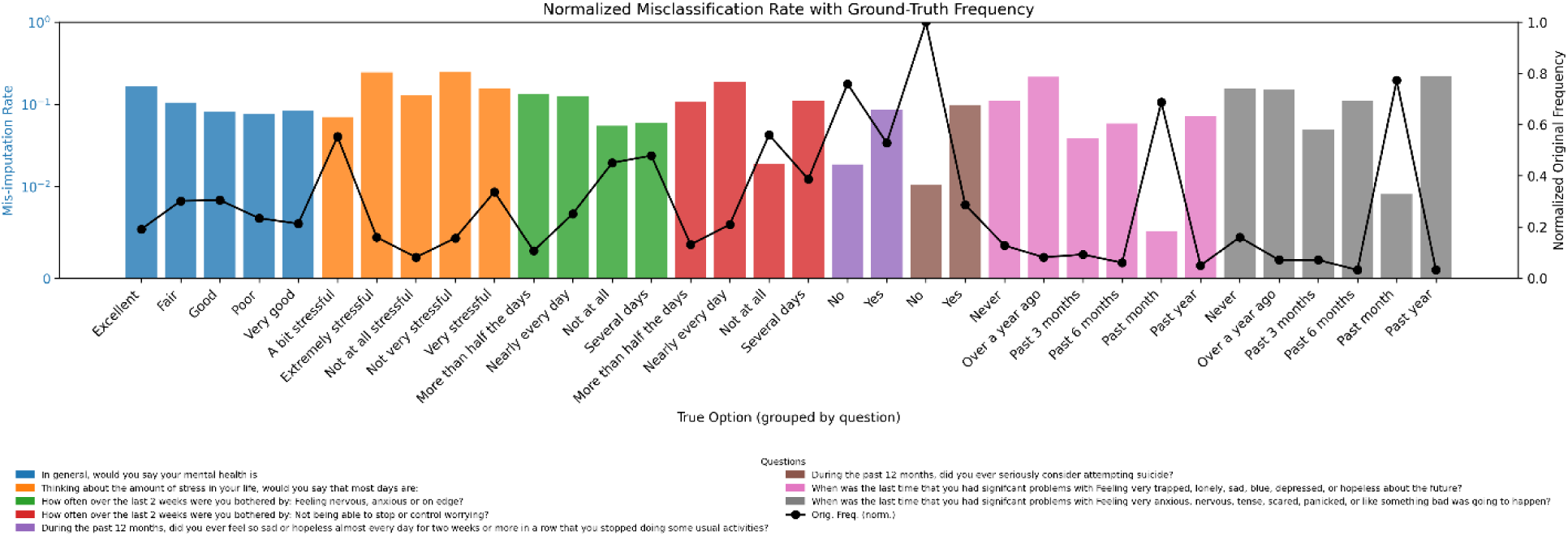
MICE Imputation misclassification distribution from the MAR generated dataset (18.32% missingness). Each bar is the frequency of misclassification for every option grouped using color for each question. The line depicts the frequency of total options from the original ground truth dataset.

**Figure 4:**
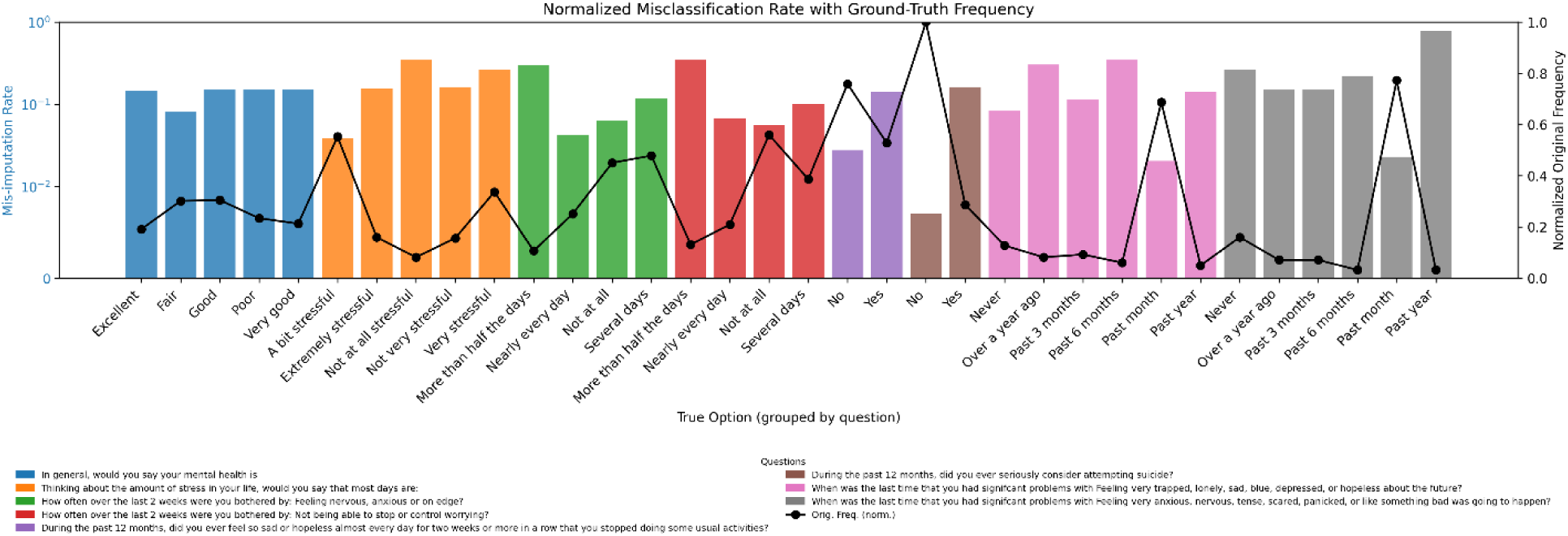
MICE Imputation misclassification distribution from the MCAR generated dataset (25% missingness). Each bar is the frequency of misclassification for every option grouped using color for each question. The line depicts the frequency of total options from the original ground truth dataset.

### 3.2. Generative AI (OpenAI o3) Imputation

For the MAR dataset (n = 363), the o3 model achieved a mean accuracy of 0.59 (SD = 0.12; range 0.44-0.73). Accuracy was lowest for “In general, would you say your mental health is” (0.44) and highest for “How often over the last 2 weeks were you bothered by: Not being able to stop or control worrying?” (0.73) (Figure 5). The mean closeness score was 0.85 (SD = 0.03; range 0.82-0.88).

**Figure 5:**
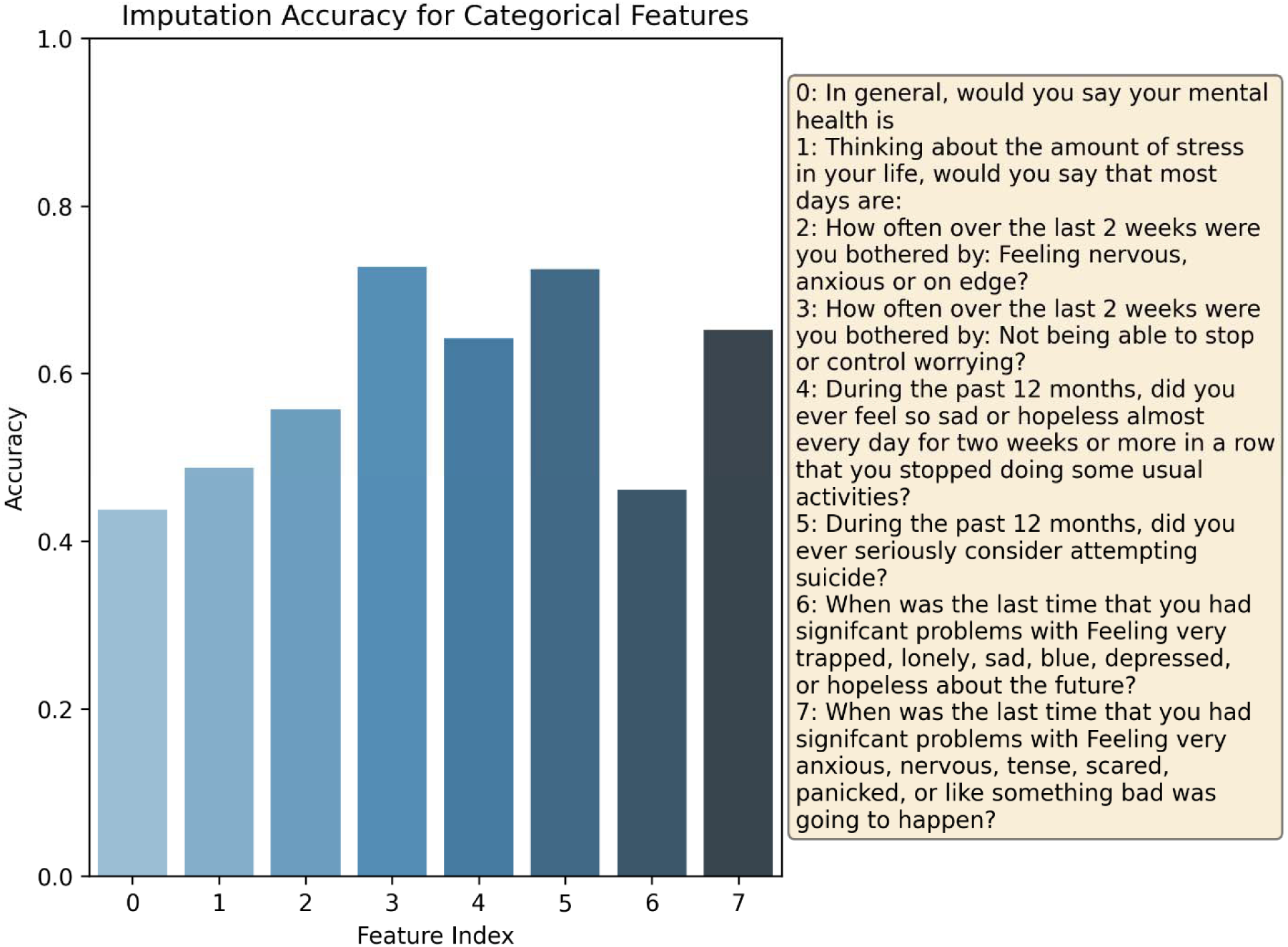
Generative AI Imputation accuracy from MAR generated dataset (18.32% missingness) using a normalized average to compute the probability of missingness. Each bar represents the accuracy of imputation compared to the ground truth dataset.

On the 25% MCAR dataset, o3 produced a mean accuracy of 0.59 (SD = 0.14; range 0.44-0.79), again lowest for “In general, would you say your mental health is” (0.44) and highest for “During the past 12 months, did you ever seriously consider attempting suicide?” (0.79) (Figure 6). The mean closeness was 0.85 (SD = 0.01; range 0.84-0.88). Unlike MICE, misclassification patterns were broadly consistent across all question types, with no clear association between response frequency and error (Figures 7-8).

**Figure 6:**
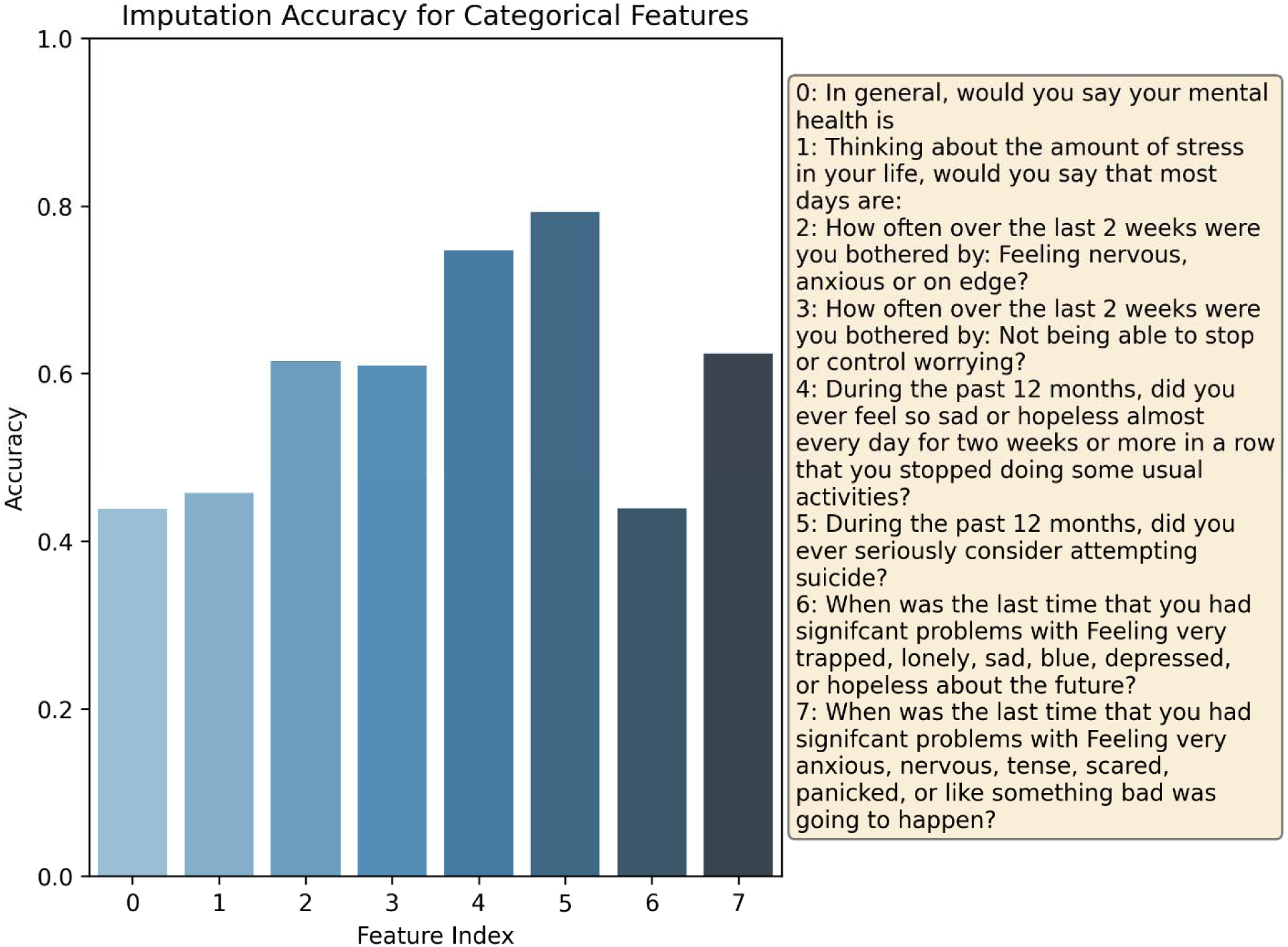
Generative AI Imputation accuracy from MCAR generated dataset (25% missingness) using a Bernoulli draw to determine missingness. Each bar represents the accuracy of imputation compared to the ground truth dataset.

**Figure 7:**
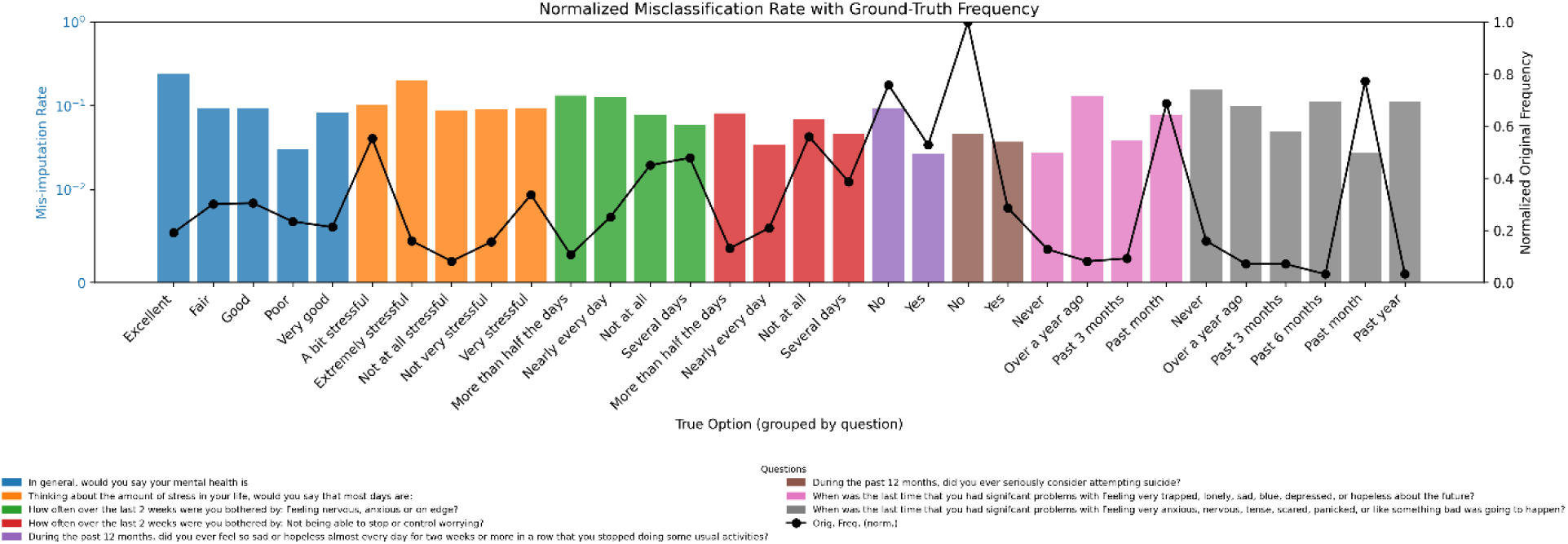
Generative AI Imputation misclassification distribution from the MAR generated dataset (18.32% missingness). Each bar is the frequency of misclassification for every option grouped using color for each question. The line depicts the frequency of total options from th original ground truth dataset.

**Figure 8:**
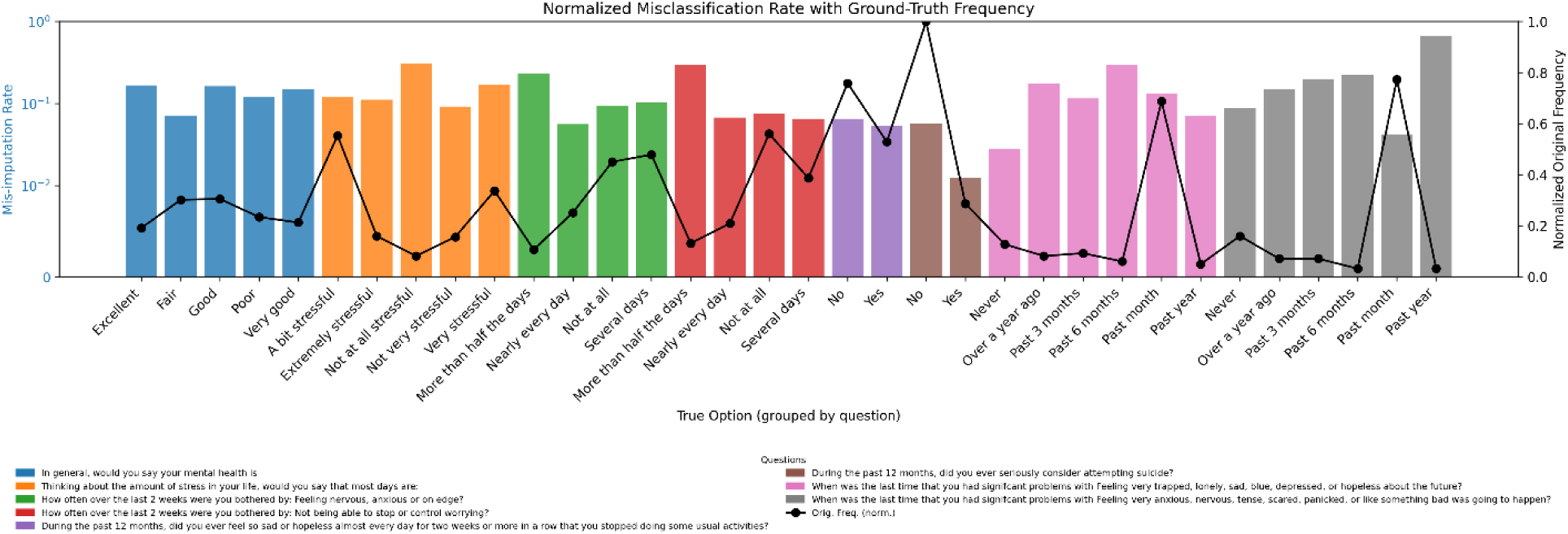
MICE Imputation misclassification distribution from the MCAR generated dataset (25% missingness). Each bar is the frequency of misclassification for every option grouped using color for each question. The line depicts the frequency of total options from the original ground truth dataset.

### 3.3. Comparative accuracy

**Table 2** summarizes average accuracies across MAR and MCAR simulations (10%, 25%, and 50%). No statistically significant differences in overall accuracy were observed between MICE and o3 (all p > 0.05).

**Table 2:** Average Imputation Accuracy and p-value with MICE imputation and OpenAI o3 imputation for each missingness simulation scenario.

| Missingness Simulation | MICE Imputation Accuracy | OpenAI o3 Imputation Accuracy | p-value |
| --- | --- | --- | --- |
| MAR | 0.60 | 0.59 | 0.655 |
| 10% MCAR | 0.59 | 0.63 | 0.318 |
| 25% MCAR | 0.59 | 0.59 | 0.917 |
| 50% MCAR | 0.53 | 0.55 | 0.665 |

## Discussion

The aim of this simulation study was to evaluate whether an LLM (OpenAI’s o3) could perform comparably to MICE for categorical data imputation in digital health dataset obtained from a digital citizen science initiative [33]. Missing data are a pervasive challenge in this domain, and while MICE has long been a standard approach [37], its reliance on predefined regression models raises concerns when applied to complex and noisy real-world data. Generative AI represents a potentially more flexible alternative, but systematic evidence on its performance in health data contexts remains scarce. This study also advances the human-centred AI agenda by demonstrating that AI-driven imputation, when grounded in digital citizen science, can integrate citizen big data and stakeholder engagement into core algorithmic processes from the conception of big data collection. The study thus demonstrates the potential feasibility of aligning LLM-based methods with human values, participatory evaluation, and digital health equity [11,12,38]. The main finding of this study is that overall imputation accuracy was similar between the o3 model and MICE across both MCAR and MAR mechanisms. The differences in classification accuracy and closeness were not statistically significant, suggesting that – at least under the tested conditions – both methods yielded comparable reconstructed datasets. This finding demonstrates that an LLM, operating in a zero-shot setting without fine-tuning, can achieve performance on par with a widely used statistical standard.

Closer consideration of the results, however, highlights important nuances. First, while both methods maintained broadly consistent performance across low to moderate levels of missingness (10–25%), a drop in performance was observed at higher levels of data loss. For the AI model, accuracy declined more steeply when missingness reached 50% under MCAR conditions, indicating greater sensitivity to extreme information loss [39]. MICE, by contrast, exhibited a more gradual reduction in accuracy, reflecting the stabilizing influence of its iterative regression-based structure. This indicates that even when methods perform similarly at lower levels of data loss (10–25%), their weaknesses can become clearer when missingness is severe [40].

The consistency of results across missingness mechanisms also warrants attention. Under MAR, both approaches produced similar imputations with stable performance even as missingness increased, suggesting that both were able to leverage the available covariate structure effectively [41]. Under MCAR, however, where no auxiliary patterns were available to guide reconstruction, the AI model showed more variability at the highest level of missingness. This divergence indicates that while large language models may capture complex dependencies when structure is present, they may be less robust when such structure is absent and large portions of data are missing [42]. Future studies should further investigate this interaction between missingness mechanisms and AI performance, particularly in high-dimensional, real-world health data where MAR is more typical but MCAR cannot be ruled out.

When situated within the broader literature, these findings align with emerging reports that large language models can approximate or even match traditional statistical techniques in structured-data reasoning tasks [30,43]. Prior exploratory work has suggested that generative models are capable of recognizing dependencies across tabular variables when formatted appropriately, even without explicit statistical assumptions [44]. However, most studies to date have been domain-agnostic or focused on benchmark tasks outside of health. The present study extends this evidence base into digital health research, providing one of the first controlled comparisons of AI- and MICE-based imputation in a simulation aligned with health survey data.

The methodological implications of these findings are significant. For researchers, the demonstration of equivalence suggests that large language models could be considered as an alternative imputation strategy, particularly in situations where traditional model specification is challenging [45]. Unlike MICE, which requires careful decisions about predictor inclusion and model form, AI-based imputation relies on general reasoning capacity and contextual inference, potentially lowering technical barriers [45]. At the same time, the greater instability observed under extreme MCAR conditions indicates that reliance on AI alone may not be advisable in settings with extensive random missingness. While MICE and the AI model were tested independently, hybrid approaches that integrate the strengths of both may ultimately offer more robust solutions [46]. For instance, ensemble frameworks where AI-generated imputations are cross-validated against statistical models could mitigate biases unique to each method while improving transparency [47].

From a practical perspective, the fact that both methods produced similar reconstructions also raises questions of efficiency, scalability, and interpretability. MICE, while well-established, can be computationally intensive in high-dimensional datasets, and requires statistical expertise to implement correctly [48]. AI models, by contrast, can generate imputations rapidly without significant statistical expertise, but raise concerns about reproducibility, transparency, and potential bias introduced by their training data i.e., potentially more accessible and generalizable across noisy and complex data sets, yet can introduce bias when missing data increases. Balancing these trade-offs will be critical if AI approaches are to be integrated into mainstream health systems and digital health research workflows. Future studies should evaluate not only accuracy but also reproducibility across prompts, consistency across model versions, and transparency of reasoning processes.

While AI tools are increasingly applied in health data analysis, a critical gap remains in embedding human-centred approaches that prioritize equity, transparency, and stakeholder participation in their design and deployment [49]. And, unlike most human-centred AI studies that focus primarily on consultation during model design, very few have explored how digital citizen science can serve as large-scale, ethical surveillance through citizen-owned devices. Embedding such approaches at the beginning of data collection not only minimizes burden but also generates ethically sourced, continuous big data. These data can then be amplified by LLMs for imputation, simulation, and prevention of poor health outcomes, positioning digital citizen science as one of the foundations of human-centred AI in a digital age where ubiquitous device ownership is near universal. Thus, beyond methodological comparability, our study highlights how digital citizen science establishes a foundation for embedding human-centred AI upstream in data collection. By leveraging citizen-owned digital tools, researchers can create ethically governed, participatory ecosystems that generate continuous data streams. These data streams are not only suitable for LLM-based imputation but also for generating synthetic datasets, thereby advancing equitable prediction and prevention [7,8].

Another consideration relates to the ethical and human-centered dimensions of AI adoption in health data science. While this study focused on technical accuracy, future research must address issues of fairness, equity, and accountability. Generative models trained on broad internet-scale data may encode biases that are not immediately apparent in simulated settings but could have downstream consequences when applied to diverse patient populations [50]. Embedding human-centered AI principles into the design, evaluation, and deployment of imputation methods is therefore essential. This includes participatory evaluation with stakeholders, explicit fairness audits, and governance frameworks that ensure AI-driven imputations do not exacerbate existing inequities in health data [51].

### Strengths and limitations

This study has several notable strengths. By directly comparing a widely established statistical method (MICE) with a generative large language model in a controlled simulation framework, it provides one of the first systematic assessments of AI-based imputation in digital health contexts. The use of both MAR and MCAR mechanisms across multiple missingness levels ensured that the findings were not tied to a single data-loss scenario, enhancing the robustness of conclusions. The evaluation also employed two complementary metrics—classification accuracy and closeness—which allowed performance to be assessed not only in absolute categorical terms but also in terms of ordinal proximity. Together, these features strengthen confidence in the central conclusion that the AI model and MICE achieved broadly similar accuracy under the tested conditions.

There are, however, important limitations that suggest avenues for future research. First, the study was restricted to simulated categorical data, meaning that generalizability to real-world health data remains untested. Observational health datasets often contain greater noise, non-standardized coding, and high-dimensional features that can challenge traditional regression-based imputation. Future work should therefore replicate these analyses in authentic digital health environments to determine whether the comparability observed here extends to more complex data structures. Second, the exclusive focus on categorical variables limits the scope of the findings. Continuous and longitudinal variables—common in digital phenotyping and precision health—pose different challenges for imputation, including handling temporal dependencies and preserving distributional properties. Extending comparisons to these data types will be essential to build a comprehensive evidence base. Additionally, the study evaluated a single large language model at one point in time. Given the rapid pace of AI development, reproducibility testing across different models and model versions will be necessary to assess whether observed comparability reflects a broader class property of generative AI or is model-specific.

### Conclusion

This study demonstrates that an LLM (OpenAI’s o3) can achieve comparable performance to MICE for categorical data imputation in simulated digital health datasets, with no statistically significant differences in overall accuracy. While both methods showed stable performance under moderate missingness, the AI model exhibited greater sensitivity to extreme MCAR conditions, highlighting that method-specific vulnerabilities remain. These findings suggest that generative AI can serve as a viable complement to traditional statistical techniques, though careful consideration of context, reproducibility, and ethical implications is necessary. Future work should extend these evaluations to diverse data types and real-world settings, integrate human-centered AI principles, and explore hybrid approaches that combine the strengths of statistical and AI methods to advance imputation practice in digital health research. Beyond its methodological findings, this study contributes to the broader vision of human-centred AI in digital health by operationalizing digital citizen science as a foundation for algorithmic evaluation. By embedding community participation, fairness considerations, and transparency into imputation research, it positions large language models not only as technical tools but also as vehicles for advancing equity, trust, and participatory innovation in healthcare.

## Data Availability

All data produced are available online at https://doi.org/10.6084/m9.figshare.29963786.v1

## Notes

### Competing Interest Statement

The authors have declared no competing interest.

### Funding Statement

This work was funded by the Canada Research Chairs Program

### Author Declarations

All procedures were reviewed and approved by the Research Ethics Boards of the University of Regina and the University of Saskatchewan (REB #2017-029)

